# Neuroanatomical correlates of working memory performance in Neurofibromatosis 1

**DOI:** 10.1101/2021.10.12.21264617

**Authors:** Cameron Sawyer, Jonathan Green, Ben Lim, Gorana Pobric, JeYoung Jung, Grace Vassallo, D. Gareth Evans, Charlotte J Stagg, Laura M Parkes, Stavros Stivaros, Nils Muhlert, Shruti Garg

## Abstract

**Background:** Neurofibromatosis 1 (NF1) is a single-gene neurodevelopmental disorder associated with cognitive and behavioural impairments, particularly with deficits in working memory. This study investigates the cerebral volumetric differences in adolescents with NF1 as compared to typically developing controls and how working memory task performance is associated with these differences.

**Methods:** 31 adolescents aged 11-17 years were compared to age and sex-matched controls. NF1 subjects were assessed using detailed measurement of working memory at baseline followed by a 3T MR scan. A voxel-based morphometry approach was used to estimate the total and regional gray matter volumetric differences between the NF1 and control groups. The working memory metrics were subjected to a principal component analysis (PCA) approach. Finally, we examined how the components derived from PCA correlated with the changes in gray matter volume in the NF1 group, after adjusting for age, sex and total intracranial volume.

**Results:** The NF1 cohort showed increased gray matter volumes in the thalamus, globus pallidus, caudate, putamen, dorsal midbrain and cerebellum bilaterally as compared to controls. The PCA yielded three independent behavioural components reflecting high memory load, low memory load and auditory working memory. Correlation analyses revealed that increased volume of the inferior lateral parietal cortex was associated with poorer performance on the high working memory load tasks. Increased volume of posterior cingulate cortex, a key component of the default mode network (DMN) was significantly associated with poorer performance on low working memory load tasks.

**Discussion:** This is the first study to examine the neuroanatomical correlates of working memory in NF1 adolescents. Consistent with prior literature we show larger subcortical brain volumes in in the NF1 cohort. The strong association between posterior cingulate cortex volume and performance on low memory load conditions supports previously suggested hypotheses of deficient DMN structural development, which in turn may contribute to the cognitive impairments in NF1.

## BACKGROUND

Neurofibromatosis 1 (NF1) is a common, single-gene autosomal dominant neurodevelopmental disorder with birth incidence of 1:2700 ^1^. It is caused by mutations in tumour suppressor NF1 gene, which is crucial for regulation of the RasMAPK molecular pathway. NF1 affects multiple organs but is most abundantly expressed in the central nervous system ^2^. Consequently, it is associated with a range of cognitive impairments and is frequently comorbid with disorders such as attention deficit hyperactivity disorder (ADHD) in about 40%-50% ^3, 4^ and Autism Spectrum Disorder in 25% of the paediatric NF1 population. For a substantial majority, learning is affected ^5, 6^ with impairments in all aspects of executive function ^7^ but visuospatial Working Memory impairments are considered one of the hallmark features of NF1 ^8^.

The NF1 gene encodes for neurofibromin which while ubiquitously expressed in the brain shows an enriched pattern of expression in the inhibitory interneurons ^9^. Neurofibromin is critically involved in cell cycle regulation through negative regulation of the RasMAPK pathway. Studies in *Nf1* heterozygous knockout mice have shown that dysregulation of the RasMAPK pathway alters GABAergic neurotransmission both in the hippocampus and the cortex, with resultant deficits in synaptic plasticity and learning. GABAergic inhibition is important not only for the stability of neuronal networks, but it plays a fundamental role in shaping cortical development including regulating progenitor proliferation, migration and maturation of neurons ^10^. In NF1, increased GABAergic activity disrupts plasticity during the critical periods^11^ and brain development. Indeed, macrocephaly early in development is one of the earliest and most well replicated brain findings in individuals with NF1 ^12^. Further, T2-weighted hyperintensities are commonly observed in the midbrain, brainstem and thalamus, and more rarely in the deep cerebral white matter and cortex ^13, 14^ in 70% of the pediatric population ^15^. They are thought to represent, in subcortical areas, foci of intramyelinic oedema and in the deep white matter of the cerebrum and the cerebral cortex, areas of dysplasia or hyperplasia ^13, 16^.

Given both brain structure and function are affected in NF1, several studies have tried to link anatomical changes in the NF1 brain to aspects of cognition but with mixed results. A well-replicated finding is an increase in volume of the corpus callosum which has been shown to be inversely proportional to verbal and non-verbal intelligence^17^ and to the severity of ADHD symptomatology ^18^. Increases in total brain volume, attributed to increases in either white matter or combined white and gray matter volumes ^19^, has not been found to be associated with neuropsychological function ^20^. Studies of regional brain volume changes in NF1 have revealed hypertrophy in subcortical structures (such as the thalamus and putamen) and the cerebellum ^21^, and links between left/right asymmetry of the auditory cortex (planum temporale) and language ability and academic achievement ^22^. Associations between specific gray matter alterations and cognition have not yet been established.

In this study we examined whether alterations in cortical and subcortical brain volumes in NF1 are linked to known cognitive deficits, particularly working memory. Working memory can be defined as short-term maintenance of information in the absence of sensory input. Working memory abilities play an important underlying role in acquisition of complex skills during development and strongly predict academic achievement ^23^. Along with the frontal lobes, the subcortical structures particularly the striatum support working memory function ^24^. Based on previous neuroimaging studies and an understanding of the role of Ras signaling in heterozygous Nf1 knockout animal models, our aim was to investigate (i) differences in total and local brain volumes in the NF1 cohort as compared to typically developing controls and (ii) associations between regional gray matter volumes in NF1 and performance on working memory tasks. Based on the previous literature ^21^, we predicted there would be macrocephaly within the subcortical structures particularly the lentiform nuclei.

## METHODS

### Subjects

Thirty-one adolescents aged 11-17 years were recruited via the Northern UK NF-clinical research network. Inclusion criteria included (i)Clinical diagnosis made using the National Institute of Health diagnostic criteria ^25^ and/or molecular diagnosis of NF1; (ii)No history of intracranial pathology other than asymptomatic optic pathway or other asymptomatic and untreated NF1-associated white matter lesion or glioma; (iii) No history of epilepsy or any major mental illness; (iv)No MRI contraindications. Participants on pre-existing medications such as stimulants, melatonin or selective serotonin re-uptake inhibitors were not excluded from participation.

Due to COVID-19 restrictions, it was not possible to collect study-specific control data. Instead, open-source healthy control images were used from the OpenNeuro, a neuroimaging data-sharing platform (Stanford Center for Reproducible Neuroscience, USA, 2021). Two sets of control data were used in order to age-match with our NF1 participants: images from 13 individuals (sample HCa) were taken from a study by Padmanabhan et al. 2011 (age range = 11.2-17.5, mean age = 14.8, SD = 2.27, 6 females) ^26^, and another 12 (sample HCb) were taken from a study by Geier et al. 2010 (age range = 11.7-18.5, mean age = 15.5, SD = 2.36, 5 females) ^27^.

The patient and control groups were matched for age and sex (p = 0.45 and p = 0.29 respectfully). Critically there was also no significant difference in age (p = 0.45) or sex (p = 0.41) between the two control samples (HCa and HCb).

Two patient scans were excluded following quality checking due to significant artefacts, leaving 29 images of individuals with NF1 (15 female, 14 male) available for the study. All scans from the 25 age-matched HCs were included (11 female, 14 male).

### Standard Protocol Approvals, Registrations, and Patient Consents

The study was conducted in accordance with local ethics committee approval (Ethics reference:18/NW/0762).

### Working memory assessments

For all NF1 participants detailed cognitive assessments were carried out to assess working memory. Verbal and visuospatial working memory was assessed using the n-back task. The task was programmed in-house using E-Prime software. Each participant completed verbal and visuospatial tasks at four levels of complexity- 0-back, 1-back, 2-back and 3-back tasks. For the verbal task, random letters were presented one at a time and the participant was asked to respond with a key-press if the letter corresponded to the letter one (1-back), two (2-back) or 3 (3-back) letters before. For the 0-back verbal task, participants were asked to press the key when they saw the letter ‘X’. For the visuospatial n-back task, blue squares were presented sequentially on a black 2 × 2 grid. Participants were instructed to respond with a key press if the position of the square matched the position one (1-back), two (2-back) or 3 (3-back) positions before. For the 0-back visuospatial task, participants were asked to respond with a key press when they saw an orange square. Each participant was presented with three blocks of each n-back task (24 blocks in total). All stimuli were presented for 500 ms and the inter-stimulus interval was set to 1,500 ms. Accuracy was calculated as the proportion of correctly identified hits + correct omissions within each block (correct hits + correct omissions/ total responses) averaged across each n-back condition.

Memory span was assessed using a computerised Corsi block task on the Psychology Experiment Building Language (PEBL) ^28^. In this task, 9 identical blue blocks are presented on the screen. These blocks light up on the screen in a sequence, which starts off as a simple sequence of two blocks and increases in complexity based on participant performance. The participant is asked to repeat the sequence observed on the screen. A measure of the memory span was computed. Auditory working memory was assessed using the digit span forward and back task of the Wechslers Abbreviated Scale of Intelligence ^29^.

Parent-rated Vineland Adaptive Behaviour Scale - third edition ^30^ was administered to the parents to assess child adaptive behaviour with overall functioning computed as standardized age equivalent and expressed as an Adaptive Behaviour Composite (ABC). Conners 3 rating scale^31^ was used as a standardized measure for parent reported ADHD symptoms. It consists of 27 items each rated on a 4-point Likert scale (0 = not true at all to 3 = very much true) in five subscales: attention, hyperactivity, learning problems, oppositionality and peer problems. The inattention and hyperactivity subscales are reported below.

### Principal Component Analysis

Given detailed assessment of working memory, we used the principal component analyses (PCA) approach to boost statistical reliability by combining results from multiple tests and to avoid the problems of collinearity when undertaking brain region-symptom correlations. Such an approach has been used in previous stroke related neuroimaging studies ^32^. The working memory metrics including n-back task accuracy (0,1,2 and 3 back) on the verbal & visuospatial tasks, corsi block memory span, digit span forward and backward were entered into the PCA with varimax rotation. Factors with an eigenvalue greater than 1.0 were extracted and rotated. Following orthogonal rotation, the loadings of each test allowed a clear behavioural interpretation of each factor. Individual participant scores on each extracted factor were used as behavioural covariates in the neuroimaging analyses.

### MRI data acquisition and pre-processing

NF1 patient MRI data was acquired on a Philips Achieva 3T MRI scanner (Best, NL) using a 32-channel receive-only head coil. 3D T1-weighted MRI were acquired sagittally with a magnetization-prepared rapid acquisition gradient-echo sequence (MPRAGE; TR = 8.4 ms; TE = 3.77 ms; flip angle = 8°, inversion time = 1150 ms, 0.94 mm in-plane resolution and 150 slices of 1mm).

Control data from HCa and HCb studies were acquired on a 3T Siemans Allegra scanne at Brain Research Centre, University of Pittsburgh. Structural data were acquired using sagittal 3D magnetization prepared rapid gradient echo (MPRAGE) (HCa ^26^: TR=1.5s, TE=25ms, flip angle=70°, 224 slices of 0.7825 mm; HCb ^27^: Tr=1.5s, TE=25ms, flip angle=70°, 192 slices of 1mm).

### VBM image processing

Images were segmented using SPM12 v7771 (Wellcome Trust Centre for Neuroimaging, University College London, UK) through MATLAB vR2020b (The MathWorks, Natick, MA, USA) into gray matter, white matter and cerebrospinal fluid tissue classes using unified segmentation. We then used the diffeomorphic anatomical registration using exponentiated lie-algebra (DARTEL) registration method to normalise images and generate a unique gray matter template from the full study population (i.e. patients and controls). This nonlinear warping technique minimises inter-individual variation in neuroanatomy. All scans were checked following each stage of preprocessing for quality control. The final voxel resolution was 1.5 × 1.5 × 1.5 mm. Spatially normalised images were modulated by the determinants of the Jacobian so that voxel intensities represent the amount of deformation needed to normalise the images. Finally, all gray matter segmented images were smoothed with a 8mm full-width half maximum isotropic Gaussian smoothing kernel.

### VBM statistical analysis

Voxel-based regression analysis (based on the GLM) was carried out using SPM12 using gray matter volume as the dependent variable. Age and sex were used as covariates of no interest (known to influence brain volume). Total intracranial volume was also calculated by summing the volumes of gray matter, white matter and cerebrospinal fluid using the ‘get_totals’ function in SPM12 and added as a global measure for proportional global scaling (in order to account for variation in head size)^33^.

For visualisation purposes, all thresholded SPM maps were imported into MRIcroGL (https://www.nitrc.org/projects/mricrogl) and overlaid onto the automated anatomical labelling (AAL) atlas ^34^.

Statistical analyses were carried out in the following stages:

1. We first analysed group differences in regional gray matter volumes between the NF1 and healthy control participants, after accounting for age, sex and total ICV.
2. Following from past literature we tested the hypothesis that putamen and globus pallidus would show group differences in volume. We created a bilateral ‘lentiform nuclei’ region of interest mask by combining left and right putamen and globus pallidus masks from the ‘Harvard-Oxford subcortical atlas’. We then used marsbar^35^ to extract gray matter volumes within this region, which were compared between the NF1 and typically developing controls.
3. We then analysed gray matter regions that show an association between increased volume and worse cognitive performance on each of the PCA factors in NF1 patients only, after adjusting for age, sex and ICV.
4. We then looked at whether gray matter volumes within the ‘lentiform nuclei’ mask from point 2 above were associated with PCA factors

For points 1 & 3 above we used Family-wise error (FWE) of p= 0.05 to correct for multiple comparisons. For point 3 we also looked for any other regions throughout the brain that showed associations with cognitive performance at a more lenient threshold (p<0.001, minimum cluster size = 10 voxels, uncorrected). For point 2 we assessed a single group difference between patients and controls (p < 0.05), and for point 4 we controlled for the three PCA factor associations using Bonferonni correction (i.e. p < 0.017)

## RESULTS

Clinical characteristics of the NF1 sample are presented in Table 1. There was a pre-existing diagnosis of ADHD in 8 participants and ASD in 3 participants. The overall Vineland adaptive functioning score was 68, well below the normative mean of 100. Parent-rated Conners-3 rating scale indicated significant impairments with T-scores of 78 and 69 on the inattention and hyperactivity subscales respectively.

**Table 1:** Clinical characteristics of the NF1 group

|  |  |
| --- | --- |
| **Males** | 15/31 |
| **Age (mean)** | 14.7 yrs ( 11.4-18.3) |
| **Pre-existing diagnoses(n)** |  |
| *ADHD* | 8 |
| *ASD* | 3 |
| **Medications** |  |
| *Stimulants* | 6 |
| *Atomoxetine* | 1 |
| **Vineland Adaptive Behavior** | 68.4 (13.0) |
| **Composite(mean,sd)** |  |
| **Conners (mean,sd)** |  |
| *Inattention T score* | 78.7 (13.0) |
| *Hyperactivity T score* | 69.1(18.2) |

We first compared differences in gray matter volumes between the patients with NF1 and healthy controls. The NF1 group showed significantly larger volumes in 3 clusters – the first large cluster included a large portion of medial subcortical structures including the thalamus, globus pallidus, caudate, putamen and dorsal midbrain and cerebellum bilaterally (see Figure 1), with the global maxima in left cerebellar lobules IV-V (p<0.05 FWE corrected). There were two other smaller but still significant regions of volume change, with large NF1 volumes in the brain stem (937 voxels) and left caudate nucleus (63 voxels). In the region-of-interest analysis, NF1 patients showed significantly greater volume of the lentiform nuclei compared to healthy controls.

**Fig 1:**
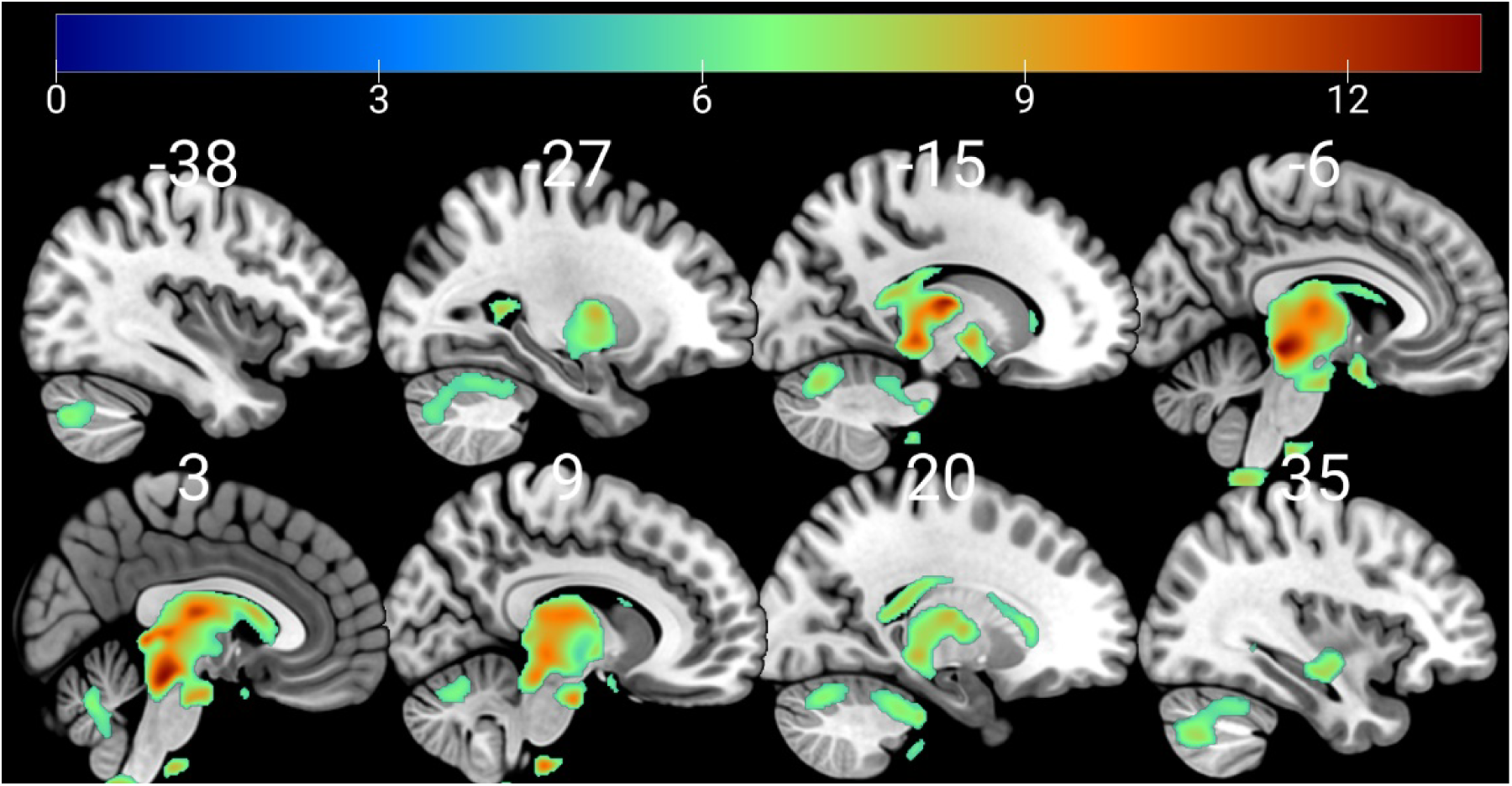
Sagittal sections showing regions of increased grey matter volume in the NF1 group compared to healthy control, superimposed on the MNI152 brain. Numbers above each slice represent MNI x-coordinates. Warmer colours represent regions of greater volume difference (see heat-map scale at top for equivalent z-values).

The working memory variables were subjected to a varimax rotated PCA which produced three independent factors with eigenvalue>1, which accounted for 54% of total variance as shown in Table 2. Measures of high working memory load loaded onto factor 1 (FAC1), low working memory load loaded onto factor 2(FAC2) and auditory working memory on factor 3(FAC3).

**Table 2:**
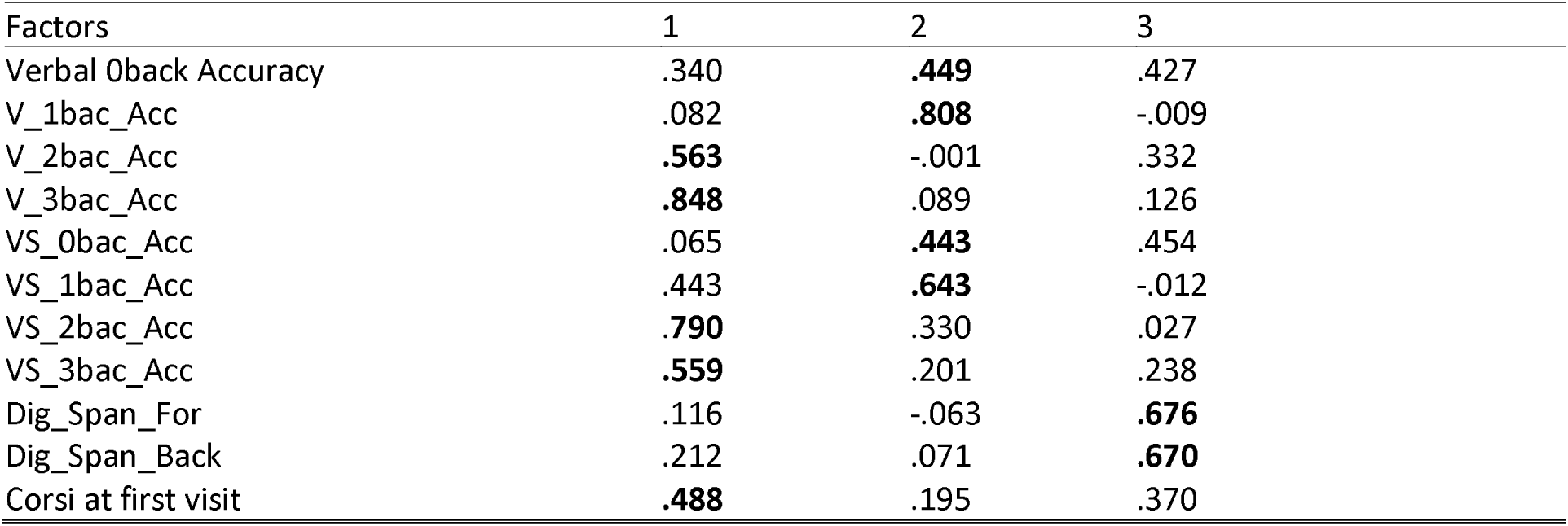
Factor loading for principal component analyses for working memory measures

Next, we examined gray matter regions where volume was associated with performance on the cognitive tasks. For this we used the first three principal components derived from the set of tasks, as described in the methods corresponding to high working memory load, low working memory load and auditory working memory. Poorer performance on high working memory load tasks was significantly associated with increased volumes in a small region of inferior lateral parietal lobe (22 voxels). Poorer performance on low working memory load was significantly associated with increased volumes in a larger region of medial gray matter centred on the posterior cingulate cortex, alongside smaller regions of lateral parietal, middle temporal and somatomotor regions (see Figure 2 and Table 3). Auditory working memory performance was associated with gray matter volumes in a single smaller region centred on the middle frontal gyrus. When focussing specifically on the region-of-interest analysis we saw no significant correlation between lentiform nuclei volumes and either FAC1 (r=-0.01, p=.98), FAC2 (r=-.15, p=.45) or FAC3 scores (r=.26, p=.17). Finally, we examined the associations between gray matter volume and age which revealed no areas that increased in volume in our NF1 sample.

**Fig 2:**
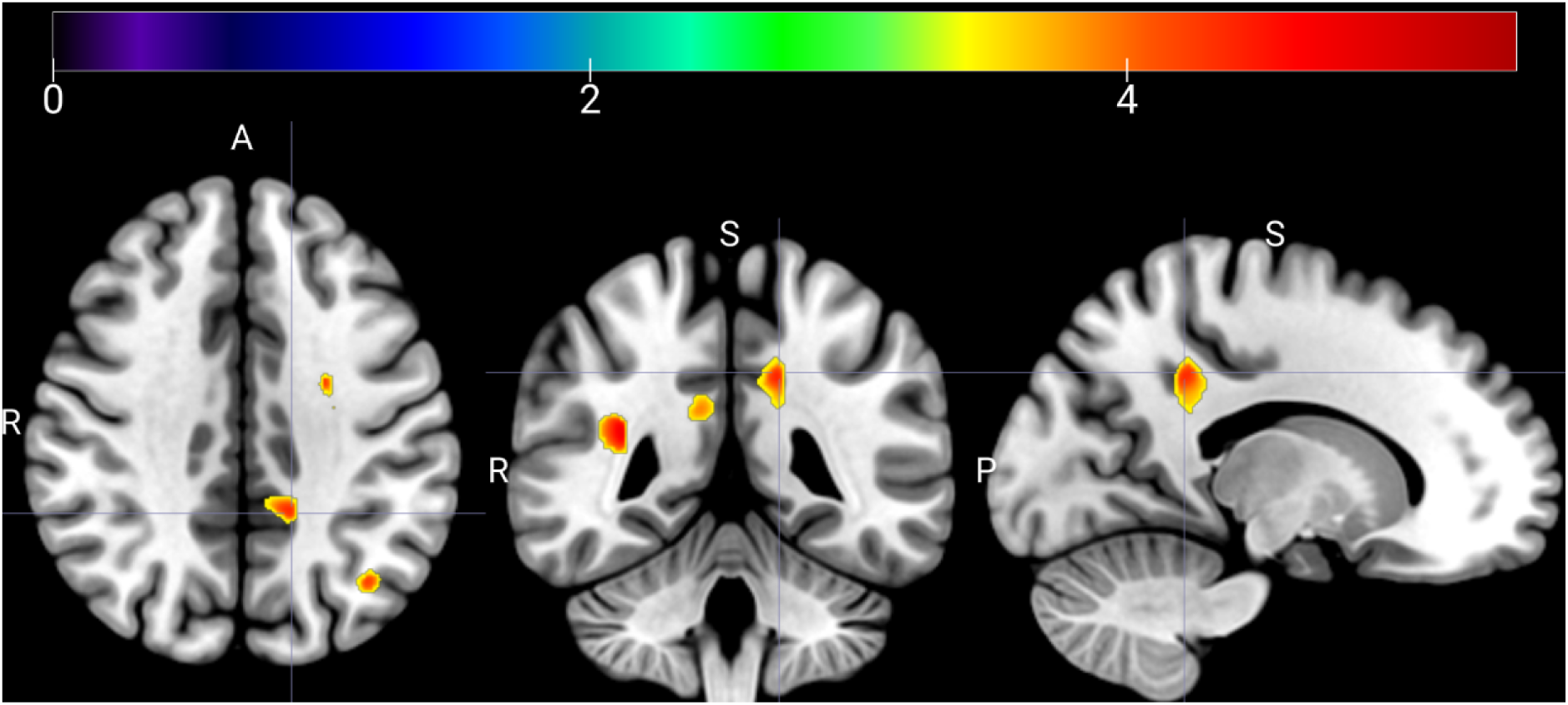
Multiplanar view of hypertrophic GM areas associated with worse low memory load capacity in adolescents with NF1. The crosshair is set at the largest voxel cluster at x = -15. Warmer colours represent regions with greatest associations between grey matter volume and FAC2 scores.

**Table 3:**
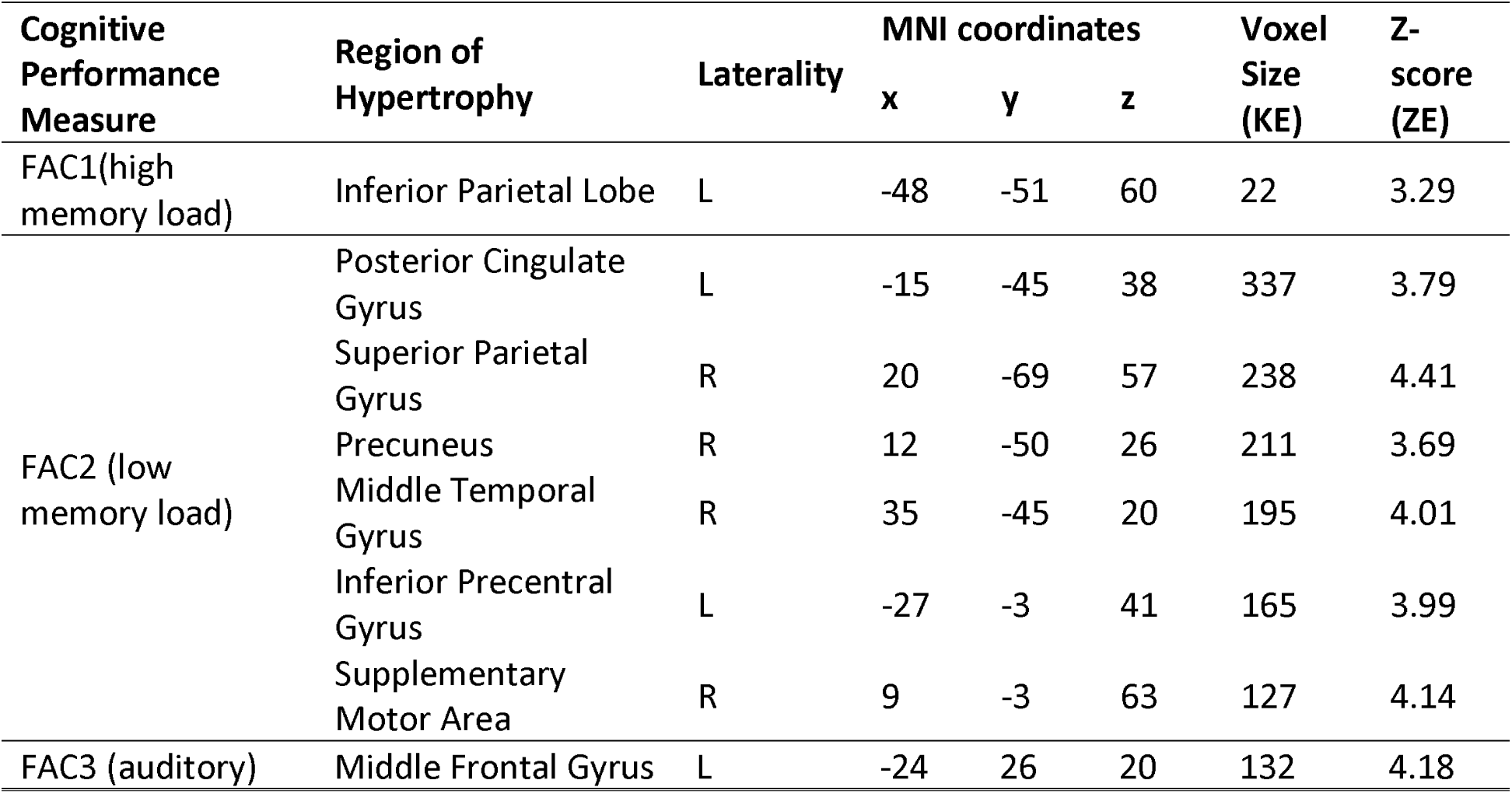
Neural correlates for principal component analyses factors

| Cognitive<br>Performance<br>Measure | Region of<br>Hypertrophy | Laterality | MNI coordinates |  |  | Voxel<br>Size<br>(KE) | Z-score<br>(ZE) |
| --- | --- | --- | --- | --- | --- | --- | --- |
|  |  |  | x | y | z |  |  |
| FAC1(high<br>memory load) | Inferior Parietal Lobe | L | -48 | -51 | 60 | 22 | 3.29 |
| FAC2 (low<br>memory load) | Posterior Cingulate<br>Gyrus | L | -15 | -45 | 38 | 337 | 3.79 |
|  | Superior Parietal<br>Gyrus | R | 20 | -69 | 57 | 238 | 4.41 |
|  | Precuneus | R | 12 | -50 | 26 | 211 | 3.69 |
|  | Middle Temporal<br>Gyrus | R | 35 | -45 | 20 | 195 | 4.01 |
|  | Inferior Precentral<br>Gyrus | L | -27 | -3 | 41 | 165 | 3.99 |
|  | Supplementary<br>Motor Area | R | 9 | -3 | 63 | 127 | 4.14 |
| FAC3 (auditory) | Middle Frontal Gyrus | L | -24 | 26 | 20 | 132 | 4.18 |

T2 hyperintensities were present in 67% (20/30) of all NF1 participants in the study. The location of the T2 hyperintensities are shown in Figure 3. The highest T2H were observed in the cerebellum, followed by the basal ganglia and the thalamus.

**Fig 3:**
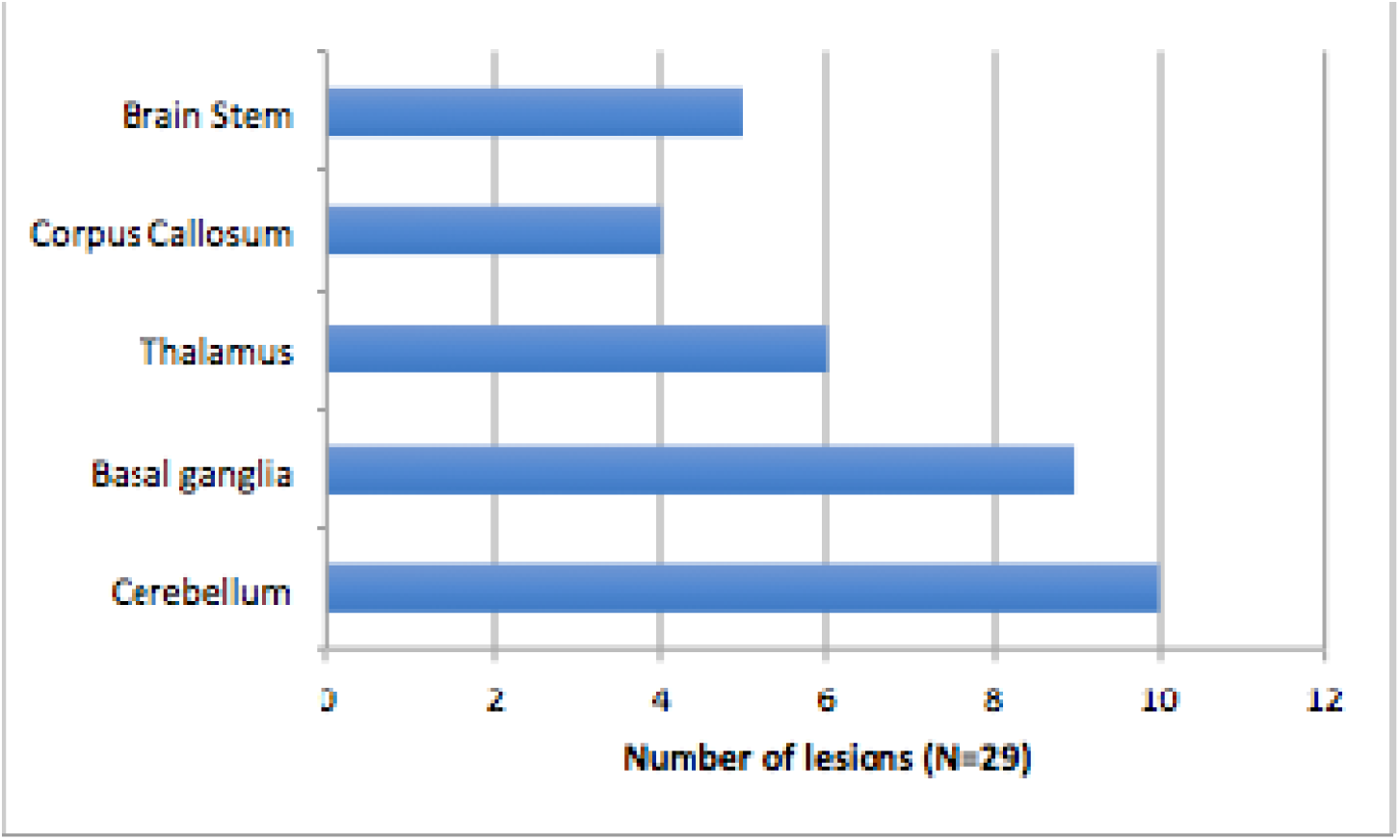
Graph showing the distribution of the T2 hyperintensities in the NF1 cohort

## DISCUSSION

To our knowledge, this is the first study to investigate the neuroanatomical correlates of working memory performance in NF1. Results confirm previously reported findings of larger subcortical brain volumes in children with NF1 as compared to controls. These included the thalamus, globus pallidus, caudate, putamen, dorsal midbrain, cerebellum and brainstem which were larger in the NF1 group. Performance on the high load working memory tasks was significantly associated with increased volume of the inferior lateral parietal cortex. Low working memory load performance was significant associated with increased volume of posterior cingulate cortex, lateral parietal, middle temporal and somatomotor regions.

Sub-cortical structures including the caudate, putamen, globus pallidus and thalamus are part of a discrete, somatotopically distributed fronto-striato-thalamo-cortical circuits that are essential for working memory and other higher executive functions including inhibition, attention and impulse control – all of which may be affected in NF1. Studies investigating the volumetric changes in the paediatric ADHD population have found significantly smaller volume of these sub-cortical structures ^36^. Similarly, recent meta-analyses of morphometric studies in idiopathic ASD populations found smaller volumes of subcortical structures including the pallidum, putamen, amygdala and nucleus accumbens ^37^. In contrast, and consistent with prior work, we observe increases in the volumes of these sub-cortical structures in NF1. Given the important role for the NF1 gene in regulating cell growth and differentiation, the increased volumes may be a direct effect of gene expression ^9^. Indeed, combined histological-MRI studies have shown that pathological changes in gray matter volume are linked to alterations in neuronal density and field fractions of glial markers ^38^. Volumetric changes in these regions thus appear to have important downstream effects on cognitive and social functioning, albeit without direct links to the cognitive abnormalities observed in our cohort. Further, the volumetric changes in the cerebellum may underlie the deficits in fine and gross motor coordination, hypotonia, problems with balance and gait reported in NF1. Moreover cerebellum was a commonly associated region for T2-hyperintensities observed in 67% of all participants, consistent with previous literature ^15^. Future research should examine the longitudinal changes in the subcortical structures.

Brain regions showing significant association with working memory performance in this study are known to be key components of the Default Mode Network (DMN). Specifically, increased volume of posterior cingulate cortex, lateral parietal and middle temporal regions were associated with poorer performance on low working memory load conditions. Core ‘nodes’ of the DMN include the posterior cingulate cortex, inferior parietal lobules, medial temporal lobe and the medial prefrontal cortex. Dynamic control of the DMN activity appears to be important for efficient cognitive function; functional MRI (fMRI) studies show a pattern of increased brain activation at rest and reduced activation during attentionally demanding tasks in these DMN regions ^39^. Although fMRI studies in NF1 are limited, two previous task fMRI studies suggest possible failure of DMN deactivation during performance of a cognitive task, suggesting increased attentional lapses during performance of cognitive tasks may be reflected in neural activity within this region ^40, 41^. In contrast, resting state fMRI studies show reduced typical long-range connectivity between the posterior cingulate and medial prefrontal cortex-both hubs of the DMN ^42, 43^ It is possible that the association between gray matter volumes in the DMN regions and cognition found in this study reflect the DMN network abnormalities that may, at least in part, underpin the cognitive impairments observed in NF1.

The posterior cingulate cortex is a highly connected, metabolically active brain region, known to be involved in self-referential processes including stimulus independent thought, day-dreaming and autobiographic memory ^44^. The posterior cingulate is particularly relevant for dynamic reallocation of visuospatial attention-needed for working memory task performance ^44^. A previous fMRI study in an ADHD population noted reduced activation of the posterior prefrontal cortex during inhibition failure, suggestive of inefficient anticipatory attention reallocation^45^. Consistent with our results of a strong association of posterior cingulate cortex with working memory performance, studies in ADHD populations also show larger gray matter volumes of posterior cingulate cortex ^36^. Taken together, posterior cingulate cortex appears to be an important DMN region mediating the NF1 associated risk of working memory impairments and ADHD symptomatology.

Our study is not without limitations. Due to a lack of local healthy control subjects, we used control data from openly available MRI datasets. We did however ensure that these datasets were matched for age and sex to our patients – in addition these data were only used for patient-control comparisons and the findings are in line with past work. Also, this work is cross-sectional and so we cannot determine the ages at which hypertrophy is most accelerated. However, we found no associations between ageing and increased volume in our NF1 sample, suggesting that increases may occur before adolescence. This would be useful to establish in future work, to understand the critical points of development for children with NF1. Finally, future studies could combine VBM approach with functional approaches such as functional MRI to better characterise brain structure-function relationship in NF1 ^46^.

Neuroimaging studies of the NF1 brain are important to understand the neural consequences of the NF1 mutations. As a single-gene disorder, NF1 present a valuable model to study the impact of the gene mutation on the brain and downstream on cognition and behaviour. Our study elucidates the link between structural brain abnormalities in NF1 and working memory function. We confirm previous fundings of increased volume of subcortical structures and demonstrate a distinct neural signature associated with working memory impairments in NF1. Increased gray matter volumes of the DMN brain regions are associated with poorer working memory performance. Our findings support the hypothesis of deficient DMN structural development, which may in turn contribute to the cognitive impairments in NF1. These findings offer further insights into the brain basis of cognitive impairments seen in NF1. Further functional brain imaging studies clarifying the specific mechanisms and functional impact associated with these structural differences are needed specially to inform strategies for future intervention trials.

## Data Availability

All healthy control data was taken from OpenNeuro (referenced in paper). NF1 cohort data will be made available on the Sage Bionetworks portal.

https://openneuro.org/datasets/ds000121/versions/00001

https://openneuro.org/datasets/ds000120/versions/00001

https://www.synapse.org/

## Acknowledgements

The authors thank the radiographers at the Manchester Clinical Research Facility: Neal Sherratt, Sarah Lehmann and Barry Whitnall for their help and assistance in acquiring the imaging data. The authors also wish to thank the patients and families that participated in this study. This work is supported by the Neurofibromatosis Therapeutic Acceleration Program (NTAP) through a Francis Collins Scholarship to SG. DGE is supported by the all Manchester NIHR Biomedical Research Centre (IS- BRC- 1215-20007). JG is supported by NIHR Senior Investigator Award. JJ is supported by a Beacon Anne McLaren Research Fellowship (University of Nottingham). This article is supported by an Agreement from The Johns Hopkins University School of Medicine and the Neurofibromatosis Therapeutic Acceleration Program (NTAP). Its contents are solely the responsibility of the authors and do not necessarily represent the official views of The Johns Hopkins University School of Medicine.

